# Bayesian Feature Selection for Multi-valued Treatment Comparisons: An Electronic Health Records Study of Vasopressor Effectiveness

**DOI:** 10.1101/2024.12.19.24319363

**Authors:** Yunzhe Qian, Bowen Ma

## Abstract

Analyzing treatment effectiveness from electronic health records (EHR) presents unique challenges in causal inference, particularly when comparing multiple treatment options with high-dimensional covariates. We propose a novel framework combining instrumental variable (IV) analysis with advanced Bayesian feature selection methods and neural networks to estimate causal effects in multi-valued treatment settings. Our approach addresses three key methodological challenges: handling multiple treatment comparisons simultaneously, comparing Bayesian feature selection methods, and selecting relevant features while capturing complex nonlinear relationships in outcome models.

Through extensive simulation studies, we demonstrate that spike-and-slab priors achieve superior performance in treatment effect estimation with the lowest mean absolute bias (0.071) compared to ALL (0.074), LASSO (0.080), and Bayesian LASSO (0.083) methods. The consistency of bias control across treatment pairs demonstrates the robustness of our Bayesian feature selection approach, particularly in identifying clinically relevant predictors.

We apply this framework to compare three commonly used vasopressors (norepinephrine, vasopressin, and phenylephrine) using MIMIC-IV data[1]. Using physician prescribing preferences as instruments[2, 3, 4], our analysis reveals a clear hierarchical pattern in treatment effectiveness. Vasopressin demonstrated superior effectiveness compared to both norepinephrine (ATE = 0.134, 95% CI [0.115, 0.152]) and phenylephrine (ATE = 0.173, 95% CI [0.156, 0.191]), while phenylephrine showed inferior outcomes compared to norepinephrine (ATE = -0.040, 95% CI [-0.048, -0.031]).

Our methodological framework provides a robust approach for analyzing multi-valued treatments in high-dimensional observational data, with broad applications beyond vessopressors in critical care. The integration of instrumental variable analysis, Bayesian feature selection, and advanced modeling techniques offers a promising direction for using EHR data to inform treatment decisions while addressing key challenges in causal inference.

## Introduction

The surge in electronic health record (EHR) data has created unprecedented opportunities for comparing treatment effectiveness in real-world clinical settings. However, extracting reliable causal insights from EHR data presents substantial methodological challenges, stemming from the complex interplay of unmeasured confounding mechanisms, particularly confounding by indication where treatment assignment is intrinsically linked to disease severity. These challenges are compounded by the high-dimensional nature of EHR data, featuring complex covariate structures, non-random missing data patterns, and intricate temporal dependencies. Such complications are particularly salient in critical care settings, where treatment decisions are both complex and time-sensitive, as exemplified in the selection of vasopressors for patients with shock.

While recent methodological advances have addressed individual aspects of these challenges [5, 6], a comprehensive framework for causal inference with EHR data remains elusive. We address this gap by developing an integrated three-component approach: leveraging instrumental variable (IV) analysis to address confounding by indication, implementing sophisticated Bayesian feature selection methods to identify informative covariates, and employing flexible neural networks for counterfactual prediction. Our approach extends existing IV methods, which have predominantly focused on binary treatments [7, 8, 9, 10, 11], to accommodate multiple treatment regimes. Building upon the theoretical foundations established by Imbens [12] and Imai and Van Dyk [13], we advance beyond conventional extensions such as regression adjustment [14] and inverse probability of treatment weighting [15], developing a deep neural network instrumental variable framework specifically designed for high-dimensional observational studies [16].

Our methodological framework advances the field in three key directions: (1) extending instrumental variable methods to handle multi-valued treatments in high-dimensional covariate spaces, (2) providing a systematic comparison of feature selection techniques in EHR-based causal inference settings, and (3) incorporating neural networks to capture complex treatment effect heterogeneity. The high dimensionality of EHR data, with hundreds of potential effect modifiers, necessitates robust feature selection methods. While various Bayesian approaches exist, their comparative performance in causal inference settings remains poorly understood [17]. Our work provides a comprehensive evaluation of four methodologies: Bayesian spike and slab priors, Bayesian LASSO, standard LASSO, and no feature selection, examining their impact on the bias-variance tradeoff in treatment effect estimation.

We demonstrate our framework’s utility through simulation and the analysis of the MIMIC-IV database (N=23,487)[1], focusing on the comparative effectiveness of three commonly prescribed vasopressors: norepinephrine, phenylephrine, and vasopressin. We exploit variation in physician prescribing preferences as instrumental variable [2], enabling causal effect estimation while accounting for unmeasured confounding. This comprehensive approach reveals important treatment effect heterogeneity that traditional methods fail to detect. The work extends beyond vasopressor selection in critical care. Our framework provides a generalizable methodology for analyzing treatment effectiveness using EHR data, synthesizing advanced causal inference methods and machine learning techniques. This integration enables both precise individual-level prediction and robust estimation of causal effects, addressing fundamental challenges in evidence-based medicine. By providing a rigorous methodology for analyzing real-world clinical data, this research advances both statistical methodology and clinical practice, offering a pathway to more personalized treatment decisions.

## Methods

### Instrumental Variable Analysis

Our analytical framework used physician prescribing preferences as an instrumental variable to address unmeasured confounding in vasopressor choice effects on mortality. We implement a two-stage estimation procedure [2, 18], enhanced with neural networks and Bayesian feature selection to capture complex treatment effect heterogeneity.

The instrumental variable is constructed using the Herfindahl-Hirschman Index (HHI) to quantify physician prescribing concentration [19]. For each physician *i*, we calculate 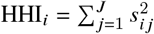, where *s*_*i j*_ represents the proportion of vasopressor *j* in physician *i*’s historical prescriptions, and *J* denotes the total number of vasopressor options. The HHI ranges from 1/*J* (equal usage) to 1 (single vasopressor usage), providing a continuous measure of prescribing preference that satisfies key IV assumptions [20].

For valid causal inference, our instrumental variable approach relies on three fundamental assumptions:

- Relevance: Physician prescribing preferences must be strongly associated with actual vasopressor choices. Formally, this requires Cov(*Z*_*i*_, *T*_*i*_ | *X*_*i*_) ≠ 0, where *Z*_*i*_ represents physician preferences, *T*_*i*_ denotes vasopressor choice, and *X*_*i*_ are observed covariates. We validate this assumption through first-stage F-statistics, requiring F > 10 to ensure strong instrument status [21]. In practice, this means physicians with a documented preference for a particular vasopressor must demonstrate significantly higher rates of prescribing that medication to their patients.
- Exclusion: Physician preferences must affect patient mortality only through their influence on vasopressor selection. This assumption requires that any impact of physician prescribing patterns on patient outcomes operates exclusively through the choice of vasopressor, not through other channels. We support this assumption by excluding physicians with fewer than 10 prior vasopressor prescriptions to ensure stable preference measures, adjusting for hospital-level factors that might influence both prescribing patterns and outcomes.
- Independence: The physician preference instrument must be independent of unmeasured confounders affecting patient outcomes. Mathematically, this requires (*Y*_*i*_ (*t*), *T*_*i*_ (𝒵)) ⊥ *Z*_*i*_ | *X*_*i*_, where Y_*i*_ (*t*) represents potential outcomes under treatment *t*. This assumption implies that physician prescribing preferences are not systematically related to unobserved patient characteristics that influence mortality risk. We validate this assumption by demonstrating balance in observed patient characteristics across physicians with different prescribing patterns and through sensitivity analyses examining the impact of potential violations.
- Additionally, for multi-valued treatments, we require the Monotonicity assumption: For any two treatments *j* and *k*, if physician A’s preference for *j* over *k* is stronger than physician B’s, then any patient who would receive treatment *j* from physician B would also receive it from physician A. Formally: *P*(*T*_*i*_ (𝒵_1_) = *j*) ≥ *P*(*T*_*i*_ (𝒵_2_) = *j*) for all 𝒵_1_ > 𝒵_2_. This assumption rules out “defier” behavior where patients systematically receive opposite treatments from what physician preferences would predict.

Our two-stage model is specified as follows:

#### Stage 1 (Treatment Model)

We employ a multinomial logistic regression to model treatment assignment:

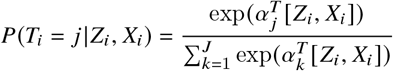

with likelihood:

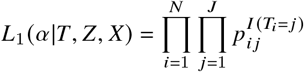

where *T*_*i*_ ∈ {1, …, *J*} represents the vasopressor choice for patient *i* among *J* possible treatments, *Z*_*i*_ is the physician preference score, *X*_*i*_ represents patient covariates, and *α*_*j*_ are the treatment-specific coefficients.

#### Stage 2 (Outcome Model)

Using the predicted treatment probabilities 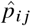 from Stage 1, we model the mortality outcome using a neural network:

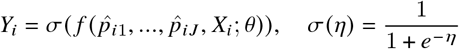

where *Y*_*i*_ is the binary mortality outcome and *f* (·) is a deep neural network with parameters *θ*. The network architecture consists of three hidden layers with ReLU activation functions and incorporates Bayesian spike-and-slab priors on the weights to enable automatic feature selection and uncertainty quantification.

### Feature Selection Methods

We implemented and compared three Bayesian and frequentist approaches for feature selection: spike-and-slab priors, Bayesian LASSO, and classical LASSO. Each method offers distinct advantages for handling high-dimensional covariate selection in our instrumental variable framework.

#### Spike-and-Slab

To perform feature selection within a Bayesian framework, we employed spike-and-slab priors on the weights of a two-layer neural network. The Spike-and-Slab approach gets its name from its two components: the spike, which drives weights of irrelevant features exactly to zero, and the slab, which allows non-zero values for significant predictors. This combination ensures both sparsity and flexibility, making it ideal for high-dimensional feature selection tasks. This method combines a discrete spike component (set at zero) and a continuous slab component, modeled by a Gaussian distribution, ensuring sparsity in the selected predictors.

The prior for each weight *θ*_*j*_ is defined as:

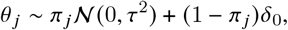

where *π*_*j*_ is the inclusion probability for weight *θ*_*j*_, *δ*_0_ is a point mass at zero (the spike component), and *τ*^2^ is the variance of the slab component, ensuring the diffuse nature of non-zero weights. The inclusion probabilities *π*_*j*_ were assigned a Beta prior:

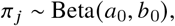

with hyperparameters *a*_0_ = 1 and *b*_0_ = 4, reflecting our prior belief that approximately 20% of the weights are non-zero. To control the global shrinkage of weights, *τ* was assigned a Half-Cauchy prior:

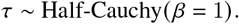

### Algorithm Implementation

The spike-and-slab model was implemented using the `PyMC5` package, a probabilistic programming library for Bayesian inference [22]. Posterior sampling was performed using the No-U-Turn Sampler (NUTS), which adaptively adjusts path lengths in Hamiltonian Monte Carlo (HMC) to ensure efficient exploration of the posterior distribution. Specifically, the posterior sampling used 4 chains and a burn-in period of 1000 iterations to facilitate convergence and reduce bias. The algorithm proceeds as follows:

#### Algorithm 1 Spike-and-Slab for Feature Selection

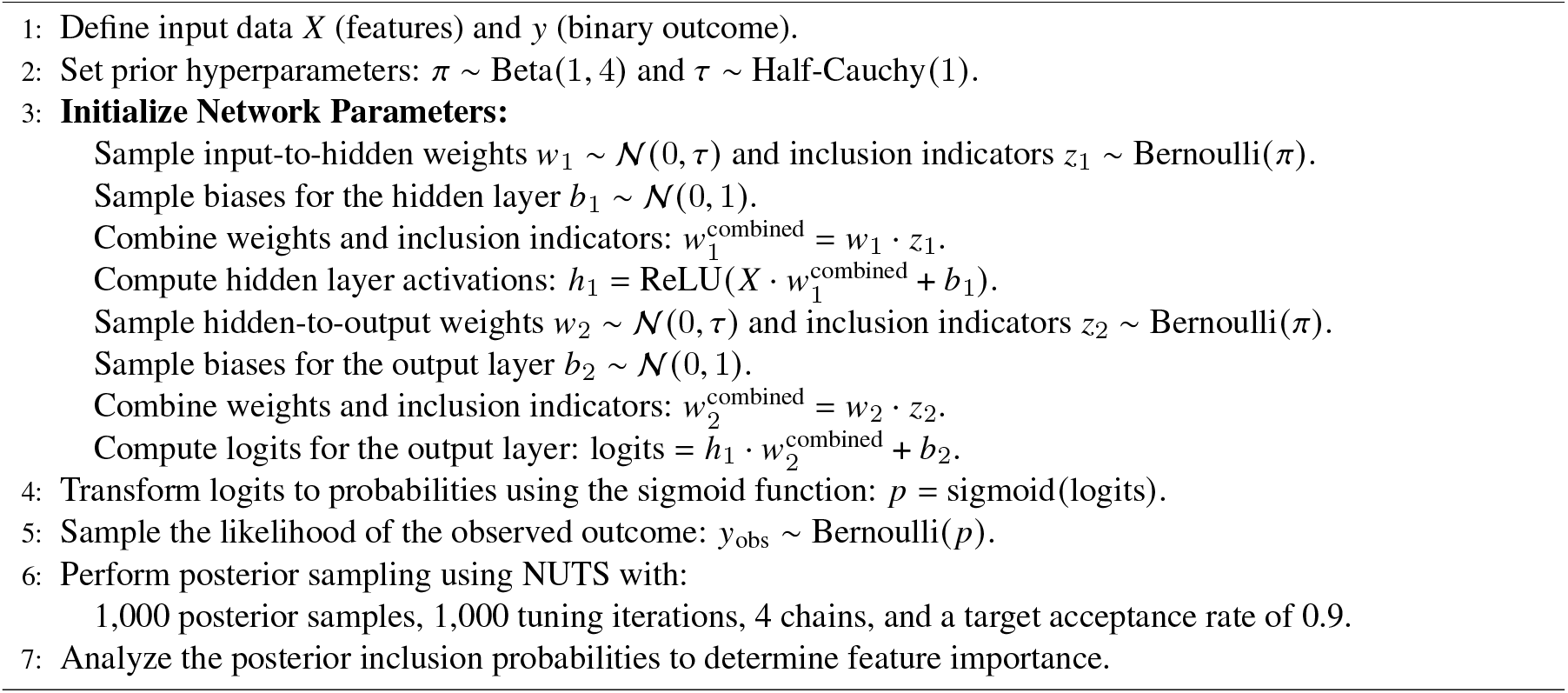

The inclusion probabilities estimated from the posterior distributions were used to identify the most informative features. Features with high posterior inclusion probabilities were retained for downstream analysis, while those with low probabilities were effectively pruned. To further quantify feature importance, we computed scores that combine both the magnitude of the weight and the probability of inclusion. Specifically, the feature importance for each predictor *j* is defined as the expected value of the absolute weight multiplied by its inclusion probability:

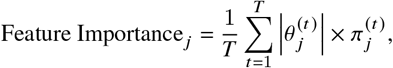

where *T* is the total number of posterior samples, 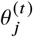 is the sampled weight, and 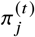 is the corresponding inclusion probability at iteration *t*. This measure reflects both the magnitude of the weight and the likelihood of the feature being included, ensuring that features with higher importance scores are identified as more influential in predicting the outcome.

#### Bayesian LASSO

To address the challenge of high-dimensional feature selection in our instrumental variable framework, we implemented the Bayesian LASSO (Least Absolute Shrinkage and Selection Operator) methodology following Park and Casella (2008) [23]. While the classical LASSO employs the double-exponential (Laplace) prior directly, this can lead to computational instability and challenges in uncertainty quantification. Instead, we adopted a hierarchical representation that ensures posterior propriety and computational stability while maintaining the selective shrinkage properties of the LASSO.

For our binary outcome setting, we specified a Bernoulli likelihood with a logit link function:

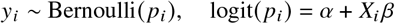

The half-Cauchy prior on the global shrinkage parameter *τ*, as recommended by Gelman (2006) and Polson (2012), provides robust regularization while maintaining sufficient posterior mass away from zero for important coefficients [24, 25]. Therefore, we implemented a hierarchical prior structure that facilitates stable computation while preserving the selective shrinkage properties of the LASSO. Specifically, we employed:

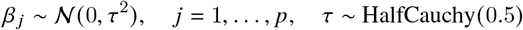

This choice represents a notable departure from the direct Laplace prior implementation, offering improved numerical stability and more reliable posterior inference.

The posterior distribution takes the form:

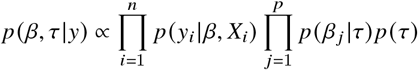

We implemented the model using `PyMC3`, employing the NUTS with 2,000 posterior samples across four chains after 1,000 burn-in iterations. Convergence was assessed through the Gelman-Rubin statistic 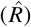 and effective sample size calculations. This representation maintains the desirable selective shrinkage properties of the classical LASSO while providing full uncertainty quantification and improved computational stability through its hierarchical structure.

Feature selection was performed using the importance score criterion that leverages the full posterior distribution of the coefficients. For each feature *j*, we computed the posterior inclusion probabilities as P(|*β*_*j*_ | > *ϵ* |data)

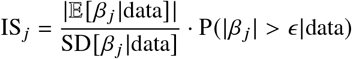

Features were retained if their importance score exceeded the 75th percentile (*ϵ*) of all scores, providing a data-driven threshold for feature inclusion that balances sensitivity and specificity while accounting for posterior uncertainty.

To ensure the reliability of our posterior inference, we implemented a comprehensive convergence diagnostic protocol. This included monitoring the Gelman-Rubin statistic 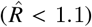 across chains, assessing effective sample sizes (minimum 400 effective samples), and visual inspection of trace plots. Additionally, we examined the energy plots from the HMC sampler to verify proper mixing and exploration of the posterior distribution.

#### Standard LASSO

Standard Lasso regression with the `TensorFlow Keras` package was used as a baseline for comparison with Bayesian feature selection techniques. The Lasso method incorporates an L1 regularization penalty, defined mathematically as:

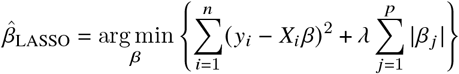

Where *λ* is the regularization parameter and *w*_*i*_ represents the weights of the model. In `Keras` implementation, the L1 penalty is applied by augmenting the loss function with a regularization term proportional to the absolute values of the model weights during optimization [26]. This mechanism facilitates feature selection by shrinking the weights of less informative features to zero, effectively removing them from the model during training. By leveraging this property, Lasso serves as a robust method for identifying important predictors while simultaneously controlling model complexity.

The training process involved setting *λ* = 1 × 10^−3^. Data preprocessing ensured that only relevant features were included and the data were split into training and testing sets. A neural network model with L1 regularization was then constructed, and hyperparameters such as the number of layers, activation functions, and optimizers were tuned to optimize performance.

### Estimation and Inference of Parameters

Parameter estimation and inference proceeds through both stages of our analysis. In the first stage, we estimate the relationship between physician prescribing preferences and treatment choices. The HHI score provides a continuous measure of physician preference, ranging from 1/*K* (equal usage of all treatments) to 1 (exclusive use of one treatment), calculated from each physician’s previous prescriptions. We assess instrument strength through F-statistics and partial *R*^2^ values, while evaluating exclusion restriction assumptions via covariate balance tests. The estimation procedure involves fitting a multinomial logistic regression model, as detailed in Algorithm 1 (lines 2-8). Parameters *α* are estimated via maximum likelihood with uncertainty quantified through bootstrapped standard errors, generating predicted probabilities 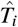 of vasopressor choice as inputs to the second stage. 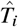 of vasopressor choice serve as inputs to the second-stage model.

In the second stage, we implement a neural network with Spike-and-Slab priors, identified as the optimal technique for feature selection, to predict binary 28-day mortality. The Spike-and-Slab approach estimates posterior inclusion probabilities (*π*_*j*_) and their uncertainties through MCMC sampling, retaining features with high inclusion probability (*π*_*j*_ > 0.5). For Bayesian LASSO, we assess convergence through Gelman-Rubin statistics 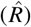 and effective sample sizes, with posterior distributions of the global shrinkage parameter *τ* and regression coefficients providing uncertainty quantification. The selected features train the final neural network model, which optimizes the binary cross-entropy loss function. Model parameters are tuned using the `Keras Tuner` framework to identify optimal hyperparameters, including optimizer choice. Treatment effect inference combines uncertainties from both stages. The average treatment effect (ATE) between treatments *k*_1_ and *k*_2_ is computed as:

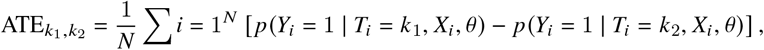

where posterior predictive distributions incorporate parameter uncertainty from both treatment and outcome models. ATEs are reported with 95% confidence intervals derived from these posterior distributions.

Model validation includes performance evaluation through standard binary classification metrics (accuracy, precision, recall, F1-score, AUC-ROC), cross-validation for assessing generalizability and mitigating overfitting, sensitivity analyses for assumption violations, and diagnostic checks of posterior distributions and MCMC convergence. Additional validation metrics include precision-recall curves, calibration plots for probability estimates, and residual diagnostics for the treatment model, ensuring robust uncertainty quantification while maintaining computational feasibility.

#### Algorithm 2 Two-Stage Estimation Procedure

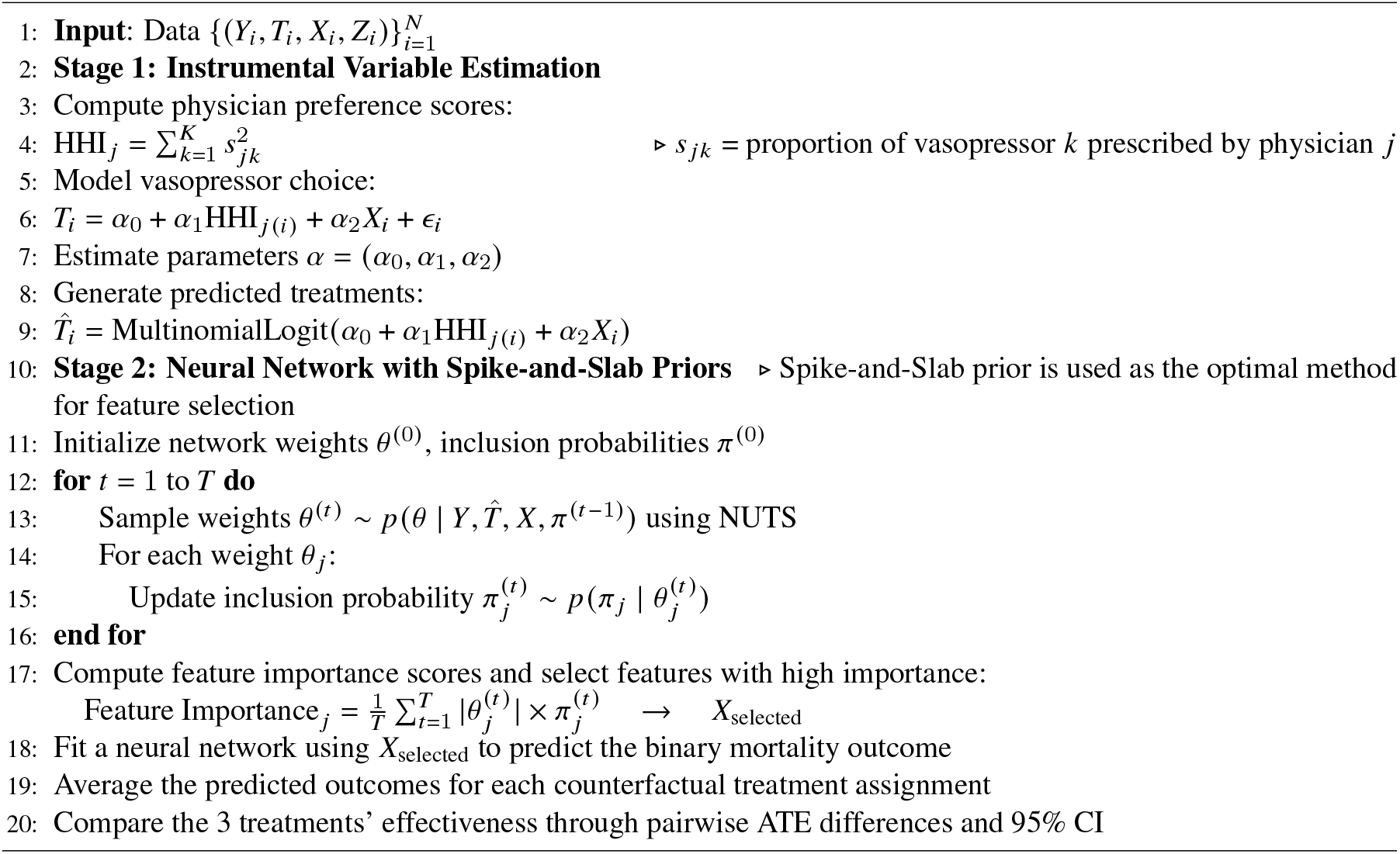

#### Model Convergence and Diagnostics Discussed

To ensure the reliability of our estimates, we conduct comprehensive convergence diagnostics and model validation. We examine trace plots of the sampled parameters to assess mixing and convergence of the Markov Chain Monte Carlo (MCMC) chains, calculating the Gelman-Rubin diagnostic statistic 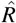 for all parameters; values close to 1 indicate convergence across chains [27, 28]. The effective sample size (ESS) for each parameter is computed to evaluate the efficiency of the sampling process, with sufficiently large ESS values indicating that the posterior samples adequately represent the true posterior distribution. Further assessment of convergence is performed through detailed evaluation of posterior distributions and trace plots to ensure stability and consistency across all chains.

During neural network training, validation accuracy is monitored to prevent overfitting, ensuring that the model generalizes well to unseen data. After training, threshold tuning is conducted to balance precision, recall, and accuracy, optimizing model performance. To evaluate the final predictive accuracy of the model, we assess precision, recall, accuracy, and the area under the receiver operating characteristic curve (AUC-ROC). Additionally, precision-recall and accuracy curves are plotted to perform threshold analysis and further validate the model’s performance.

Sensitivity analyses are typically performed by varying hyperparameters such as *τ*^2^, *a*_0_, and *b*_0_ in the Spike-and-Slab priors to assess the robustness of the results and ensure that conclusions are not unduly influenced by specific prior choices. However, due to the computationally intensive nature and long runtime of the Spike-and-Slab model, sensitivity analysis was not conducted in this study. To mitigate overfitting, Spike-and-Slab priors and dropout regularization were employed in the neural network. Training and validation loss were closely monitored during model fitting to detect potential overfitting. Furthermore, comprehensive convergence diagnostics and validation techniques [29] were implemented to ensure that the Bayesian neural network provides reliable and valid estimates for causal inference and variable selection in the context of vasopressor effectiveness.

## Result

### Numerical Studies

To evaluate our methodology, we conducted extensive simulation studies mirroring real-world observational data with multiple treatment options. The simulation framework incorporates unmeasured confounding, a valid instrumental variable, and high-dimensional covariates. For each simulation, we generated datasets with 10,000 observations. The covariates *X* ∈ ℝ^100^ were drawn from a multivariate normal distribution with zero mean and identity covariance matrix. Two unmeasured confounders (*U*_1_, *U*_2_) ∼ 𝒩 (0, 1) were introduced to influence both treatment selection and outcomes. The instrumental variable was constructed as *Z* = 0.2*U*_1_ + 0.2*U*_2_ + *ϵ, ϵ* ∼ 𝒩 (0, 1).

Treatment assignment followed an ordered logit model with six levels: *T*^∗^ = 1.5*Z* + 0.4(*U*_1_ + *U*_2_) + 0.3(*X*_1_ + *X*_2_) + *η, η* ∼ 𝒩 (0, 0.5), where final treatment *T* ∈ 1, … , 6 was determined by threshold crossings of *T*^∗^. The binary outcome Y was generated through a logistic model:

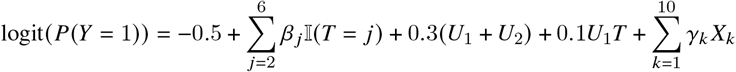

where self-defined *β*_*j*_ = (0.5, 1.5, −3.0, 2.0, 3.5) represents heterogeneous treatment effects, and self-defined *γ*_*k*_ represents the effects of the first ten covariates. The remaining covariates were included in the dataset but did not directly influence treatment or outcome, allowing evaluation of feature selection methods. The simulation design satisfies instrumental variable assumptions while maintaining realistic complexity. The instrument demonstrated adequate strength (first-stage F-statistic > 10) and appropriate exclusion restriction through its data-generating mechanism. The inclusion of both measured and unmeasured confounding, along with heterogeneous treatment effects, provides a rigorous test of our methodology’s ability to recover causal effects in challenging scenarios. All continuous variables were standardized to zero mean and unit variance to ensure stable model optimization. Since the simulated data contained no missing values, no imputation was required.

#### Feature Selection

##### Spike-and-Slab

To evaluate the convergence of the spike-and-slab model applied to the simulated dataset, we monitored the posterior distributions and trace plots of the key parameters, *π* and *τ*, as shown in Figure 1. The parameter *π* displayed strong evidence of convergence and well-mixed behavior across the four Markov chains. The trace plot for *π* shows no discernible trends, with the chains exhibiting proper mixing and stable oscillations around a consistent range of values. Furthermore, the posterior distributions of *π* align closely across all chains, indicating that the sampling process achieved reasonable acceptance probabilities. This behavior is supported by the Gelman-Rubin statistic 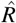 of 1.01, which is well below the threshold of 1.1, confirming that *π* converged successfully.

**Figure 1.**
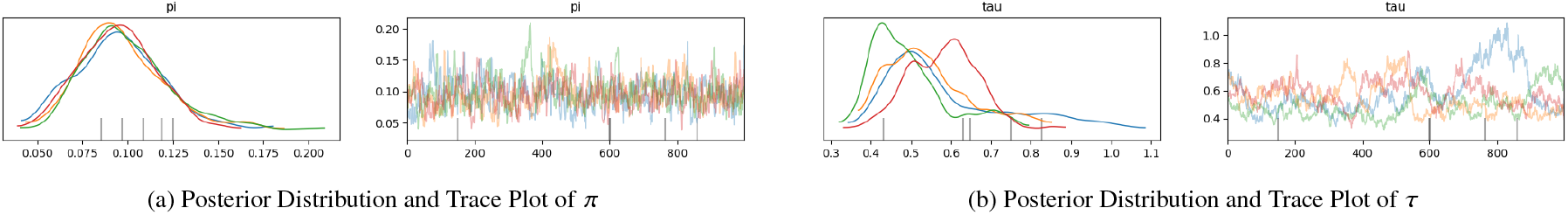
Posterior distributions and trace plots for spike-and-slab parameters *π* and *τ* on the simulated dataset.

On the other hand, the parameter *τ* demonstrated moderate convergence issues. The posterior distributions for *τ* exhibited visible misalignment across the four chains, and the trace plot revealed sustained trends with less mixing. The trajectories of the chains suggest potential issues with high acceptance probabilities, as reflected in the relatively smooth trace paths without sufficient exploration of the parameter space. While 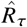 was 1.28, which is an improvement compared to the real-world analysis, it remains above the ideal threshold. These findings highlight that the estimation of *τ* remains computationally challenging, even in a controlled simulated setting, due to its sensitivity to variability in the slab component. Despite this, the runtime for posterior sampling was approximately 3 hours using 4-core parallelization, demonstrating the model’s computational feasibility for moderately sized datasets.

Given the moderate convergence issues observed in the parameter *τ*, we identified high variability in estimating the spread of significant weights, which poses challenges in determining the optimal feature set. Fine-tuning the Spike-and-Slab hyperparameters was deemed unfeasible due to computational burden discussed before. Consequently, we conducted a feature importance threshold analysis to investigate the relationship between the number of features selected by Spike-and-Slab and the accuracy of the resulting neural network model. This analysis aims to assess whether Spike-and-Slab effectively truncates noise in the dataset and identifies truly important features. Specifically, we hypothesize that reducing the feature set by removing less important features could maintain or even improve model accuracy, reflecting the robustness of the Spike-and-Slab feature selection in isolating informative predictors.

Figure 2 presents the relationship between feature importance thresholds and model accuracy. To evaluate the effectiveness of spike-and-slab feature selection, a series of neural network models were trained using features selected at varying percentiles of importance scores derived from the spike-and-slab posterior inclusion probabilities. The accuracy peaked when the bottom 15% of features were removed, meaning that the top 85% of features were retained, achieving an accuracy of 0.772. This result indicates that a large proportion of features contributed to the model’s predictive performance, and removing the least informative 15% improved accuracy while avoiding excessive sparsity.

**Figure 2.**
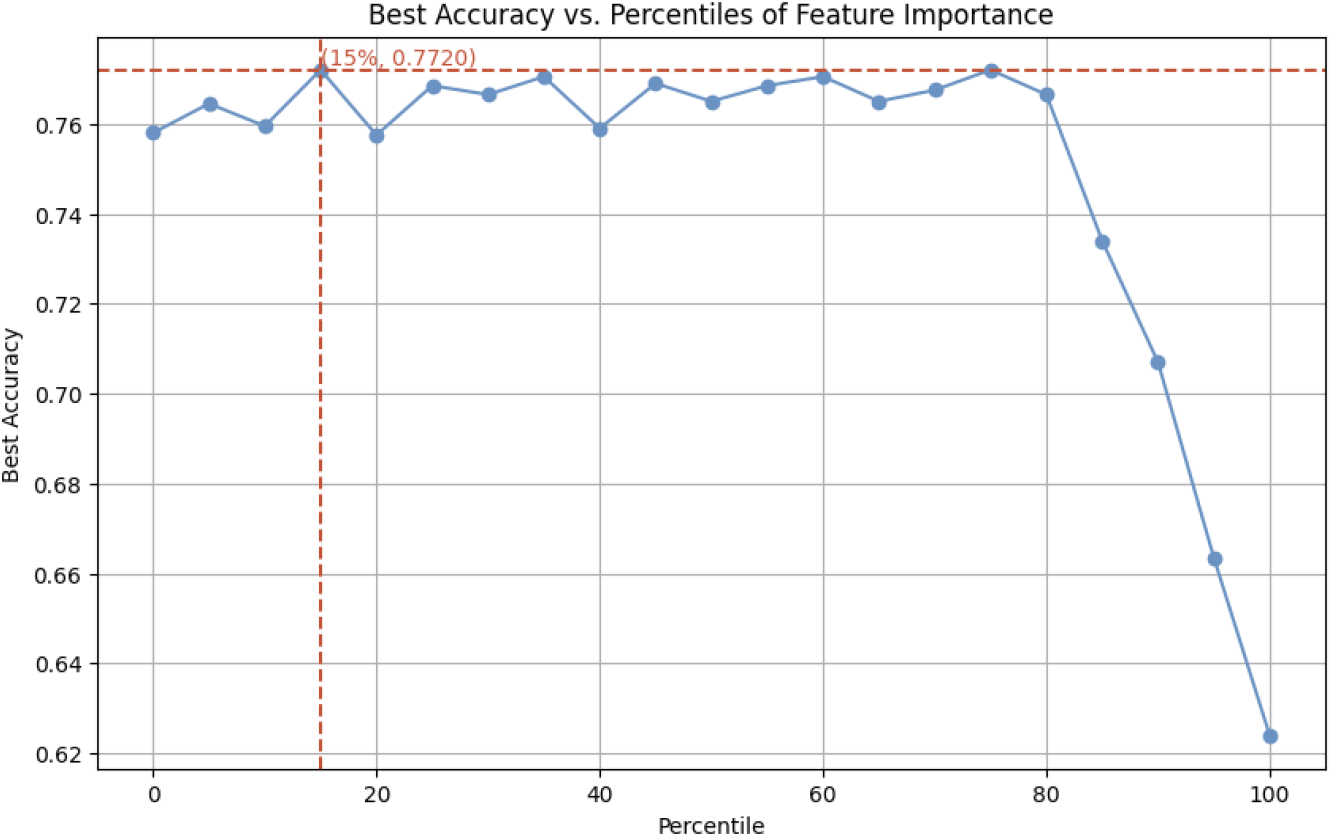
Best accuracy vs. percentiles of feature importance using spike-and-slab feature selection for the simulated dataset.

Including additional features beyond the top 85% threshold resulted in a sharp decline in accuracy, which can be attributed to the introduction of irrelevant or noisy predictors. The spike-and-slab model demonstrated its capability to perform robust feature selection under a Bayesian framework, effectively identifying informative features while discarding those with negligible contributions. While the parameter *π* achieved excellent convergence, the moderate convergence issues with *τ* highlight the challenges associated with its estimation. By removing the bottom 15% of features and retaining the top 85%, the model achieved a peak accuracy of 0.772. These results emphasize the effectiveness of spike-and-slab priors in improving predictive performance by optimizing feature selection and filtering out less informative predictors.

### Bayesian LASSO

The Bayesian LASSO demonstrated robust convergence and feature selection capabilities in our simulation study. The model achieved stable convergence across all parameters, with Gelman-Rubin statistics 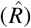 ranging from 1.000 to 1.006 for coefficient estimates. The global shrinkage parameter *τ* showed particularly strong convergence 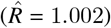, with high effective sample sizes (ESS bulk = 4623.0, ESS tail = 2781.0) indicating efficient posterior sampling. The posterior estimates demonstrated strong concentration, with *τ* estimated at 0.168 (97% HDI: [0.145, 0.192]) and intercept at 0.257 (97% HDI: [0.205, 0.308]), supported by high effective sample sizes (ESS bulk = 7133.0 for intercept) suggesting reliable posterior inference.

From the complete feature set (*p* = 112), the Bayesian LASSO identified 84 relevant predictors using the 75th percentile threshold criterion. The trace plots (Figure 3) exhibit stable mixing behavior across chains, with smooth posterior densities indicating proper convergence, while maintaining computational efficiency through parallel chain implementation. This pattern suggests a natural sparsity in the feature space, validating the LASSO’s selective shrinkage properties.

**Figure 3.**
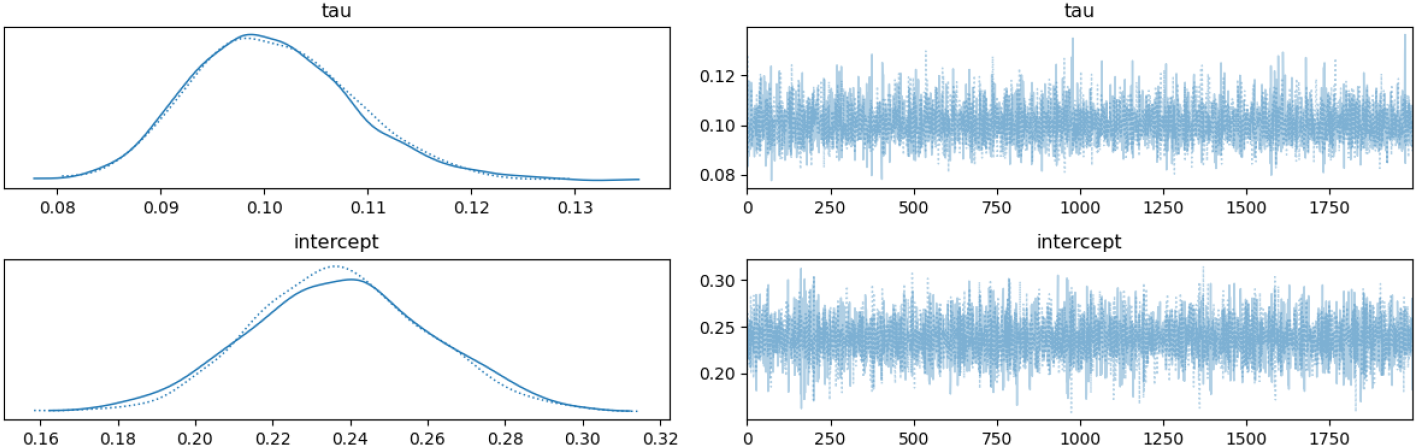
MCMC diagnostic plots for the simulation data, Bayesian LASSO model. A. Trace plots for model parameters (*τ* and intercept) showing mixing behavior across two chains. Left panels show smoothed posterior densities; right panels show detailed sampling trajectories over iterations. B. Forest plot and Posterior distributions.

### Standard LASSO

was applied to the simulated dataset to perform feature selection using *L*_1_-regularization with *λ* = 1 × 10^−3^. The *L*_1_ penalty effectively enforces sparsity by shrinking less important feature weights to zero. In this analysis, LASSO successfully shrank 2,297 out of 25,088 weights to zero, demonstrating its ability to impose sparsity at the weight level. However, no input features were completely removed, as all 112 features retained nonzero weights. This result indicates that while LASSO reduces the influence of less important features, it does not entirely eliminate any predictors in this simulated setting.

The feature importance scores, calculated as the sum of absolute weights for each feature, are illustrated in Figure 4. The feature importance distribution highlights that certain features exhibit dominant scores, contributing substantially to the model’s predictive performance, while a majority of features maintain relatively small importance values. This behavior underscores LASSO’s ability to prioritize a subset of key predictors while retaining all features.

**Figure 4.**
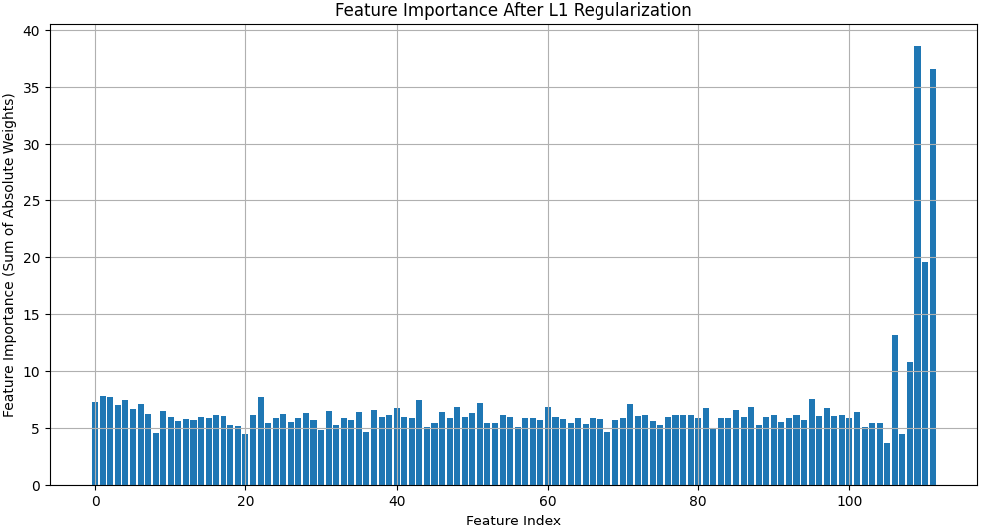
Feature importance after LASSO regularization, showing the sum of absolute weights for each input feature in the simulated dataset.

The standard LASSO model achieved a test accuracy of 0.767. While this result is competitive, it was slightly inferior to both the Bayesian LASSO model (0.771) and the spike-and-slab method (0.771). The marginal performance gap between LASSO and the Bayesian approaches reflects the limitations of LASSO’s deterministic framework, which does not account for model uncertainty or posterior distributions of the weights. In contrast, Bayesian methods can quantify uncertainty, providing a more robust framework for feature selection and predictive modeling.

These findings demonstrate the effectiveness of the *L*_1_-regularization penalty in promoting sparsity and model interpretability, even in simulated datasets. However, the superior performance of Bayesian LASSO and spike-and-slab highlights the advantages of probabilistic approaches in achieving optimal feature selection and improved predictive performance.

#### Bias

Our comprehensive bias analysis across 15 treatment pair comparisons, as illustrated in Figure 5, reveals consistently controlled bias across all estimation methods. The Spike-and-Slab method demonstrated marginally superior performance with the lowest mean absolute bias (MAB: 0.071), followed closely by ALL (MAB: 0.074), LASSO (MAB: 0.080), and BayesLASSO (MAB: 0.083). The visualization of bias estimates and their corresponding confidence intervals reveals systematic patterns across treatment comparisons, with larger biases typically observed in comparisons involving Treatment 1 (particularly T1 vs T5 and T1 vs T3). Notably, most bias estimates fall within ±0.2, with substantial overlap in confidence intervals across methods, suggesting that all approaches provide statistically comparable performance. The Spike-and-Slab method’s estimates consistently cluster closer to the zero-bias reference line. These findings indicate that while all methods achieve satisfactory bias control, the Spike-and-Slab method offers a modest but consistent advantage in estimation accuracy, particularly for certain treatment pair combinations.

**Figure 5.**
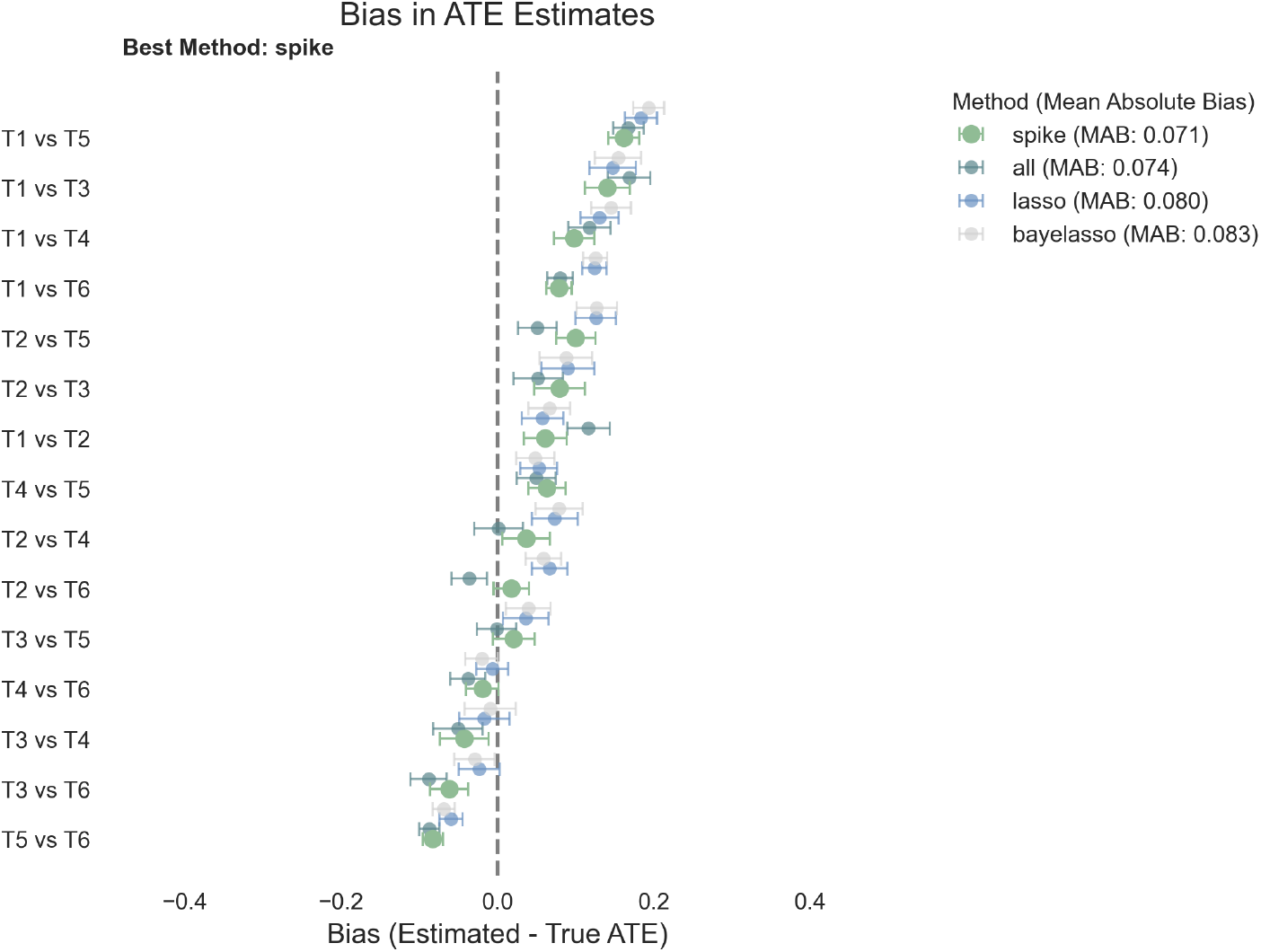
Enter Caption

### Real-World Data Application

#### Study Population and Sample Selection

We analyzed data from the MIMIC-IV critical care database (N = 23,487), employing a multi-stage sampling framework to identify eligible patients who received vasopressor therapy during ICU admission between 2008-2019. The sampling frame comprised adult ICU admissions with documented administration of norepinephrine, vasopressin, or phenylephrine. Sequential exclusion criteria were applied: age < 18 years, ICU length of stay < 24 hours, invalid vasopressor records, missing outcome data, and incomplete baseline covariates, yielding a final analytic cohort of 23,487 patients. Variable extraction followed a standardized protocol encompassing three domains: (1) high-frequency physiologic measurements (heart rate, blood pressure components, respiratory parameters, oxygen saturation) recorded at hourly intervals; (2) laboratory values (metabolic, renal, and hematologic parameters); and (3) demographic and clinical characteristics. Time-varying measurements were summarized using sufficient statistics (mean, extrema) over the initial 24-hour period. Laboratory trajectories were characterized through first and last measurements and temporal derivatives. Physiologic variables underwent range validation using established clinical thresholds, with outliers truncated at domain-specific bounds. Data preprocessing employed standardization for continuous variables (*µ* = 0, *σ;* = 1) and one-hot encoding for categorical variables to ensure scale invariance and avoid ordinal assumptions. Missing data analysis revealed 79.7% variables with missingness, with 69.84% of those showing < 3% missing values. A subset of hemodynamic and laboratory parameters exhibited higher missingness (maximum 35.75%). Simple imputation methods (mean for continuous, mode for categorical) were implemented given the low overall missingness (6.61%) and robustness of downstream feature selection procedures to imputation uncertainty. The instrumental variable construction utilized physician prescribing patterns, quantified through the Herfindahl-Hirschman Index (HHI). Inclusion required minimum prescription volume (≥ 10 patients) to ensure stable preference estimation, yielding 426 eligible physicians. The HHI provided a continuous measure of prescribing concentration (range: [0.33,1.0]), where 0.33 indicates equal distribution and 1.0 indicates exclusive use. Primary outcomes included 28-day mortality (binary) and hospital-free days (ordinal: [0,28]), with ICU length of stay as a secondary endpoint.

#### Covariate and Outcome Measurement

The measurement framework captured extensive patient-level data across multiple domains. Demographics encompassed standard variables (age, gender, race) and healthcare system factors (insurance status, admission characteristics). Clinical parameters were systematically extracted at high temporal resolution (hourly) during the initial 24 hours post-ICU admission. Cardiovascular monitoring included heart rate and multi-component blood pressure measurements (systolic, diastolic, mean arterial). Respiratory function was assessed through respiratory rate and oxygen saturation. Laboratory measurements comprised metabolic (lactate, glucose), renal (creatinine), electrolyte (potassium, sodium), and hematologic (hemoglobin, leukocytes) parameters. Each laboratory value generated six summary statistics: initial and final measurements, extrema, mean, and temporal gradient. Statistical preprocessing included removal of physiologically implausible values (< 0.1th or > 99.9th percentile) to minimize measurement error impact. Outcome ascertainment utilized multiple data sources. The primary outcome (28-day mortality) was determined through discharge disposition and hospital mortality flags. Secondary outcomes were algorithmically derived: hospital-free days (28 minus length of stay, truncated at zero for non-survivors) and length of stay (calculated from admission/discharge timestamps). The instrumental variable construction utilized unique physician identifiers linked to vasopressor orders. Prescribing preference metrics included total prescription volume, primary vasopressor choice, and two concentration measures: the HHI and a simplified concentration ratio. Additional data validation steps included temporal logic checks (e.g., administration end times posterior to start times) to ensure measurement integrity.

### Feature Selection

#### Spike-and-Slab

Convergence diagnostics for the real-world dataset revealed distinct behaviors for the parameters *π* and *τ*. As shown in Figure 6, the posterior distribution and trace plots of *π* demonstrated strong convergence, characterized by stable mixing across chains and a Gelman-Rubin statistic 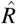 of 1.02. This indicates that *π* achieved proper mixing and convergence to a consistent region of values. In contrast, the parameter *τ* exhibited significant convergence challenges. The posterior distributions of *τ* displayed discrepancies across chains, and the trace plots revealed poor mixing with sustained trends, reflecting instability in parameter estimation. The Gelman-Rubin statistic for *τ* was 1.73, underscoring the difficulty in achieving convergence due to the high variability associated with the variance parameter. These findings suggest that the model struggles to differentiate between small but meaningful features and random noise when applied to real-world data. Additionally, the computational burden of spike-and-slab sampling was substantial, with each iteration requiring approximately 28 hours without parallelization and 7–8 hours using 4-core parallelization, making fine-tuning of *τ* computationally infeasible.

**Figure 6.**
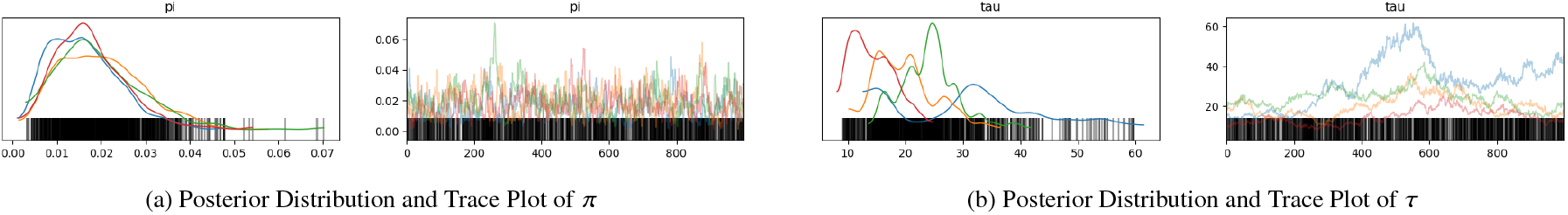
Posterior distributions and trace plots for *π* and *τ* showing convergence for *π* and poor mixing for *τ*.

Similar to the simulation framework, these convergence challenges necessitated a feature importance threshold analysis. This analysis seeks to examine the relationship between the number of selected features and model accuracy and assess whether the spike-and-slab model effectively filters noise while retaining truly informative predictors. By exploring the predictive performance of models trained with varying thresholds of selected features, we aim to determine whether reducing the feature set improves accuracy by removing irrelevant variables.

To evaluate the optimal threshold for feature selection, a neural network was trained for each possible percentile from 0 to 100, with 0 percentile using only treatment probabilities as input features and 100 percentile including all features selected by spike-and-slab. The purpose of this analysis was to demonstrate the ability of spike-and-slab to filter out non-informative features and noise. By reducing the number of input features, the neural network was able to achieve improved prediction results. As shown in Figure 7, the highest accuracy (0.822) was achieved when using the top 75% of features ranked by spike-and-slab. This result highlights the effectiveness of the spike-and-slab method in identifying informative predictors while excluding noise, enabling the neural network to yield better predictions with fewer input features.

**Figure 7.**
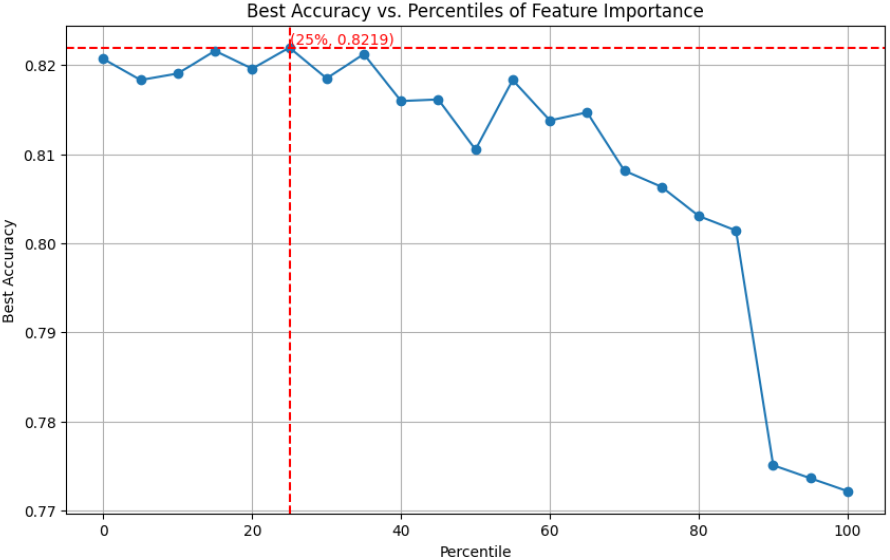
Best accuracy vs. percentiles of feature importance, showing the highest accuracy achieved with the top 75% of features.

The mechanism for deriving feature importance involved extracting posterior samples of weights and inclusion probabilities, averaging over hidden units and samples to compute scores. Features were ranked based on these scores, and the top 75% were selected after excluding redundant or irrelevant predictors. The selected features were used in downstream modeling to optimize predictive performance.

#### Computational and Convergence Challenges

Building on these methodological considerations, a key challenge lies in the computational burden of the Spike-and-Slab method, which requires several hours per iteration even with parallelization, limiting its scalability for larger datasets. The computational inefficiency of the Spike-and-Slab method can be attributed to the centered parameterization of the variance parameter *τ*, where *θ* ∼ 𝒩 (0, *τ*^2^) and *τ* follows a half-Cauchy distribution. This parameterization creates a posterior geometry known as Neal’s funnel[30]. The geometry is characterized by a narrow region near the origin, corresponding to small values of *τ*, and a wider region for larger values. This funnel restricts the movement of MCMC steps, making it difficult for the sampler to efficiently explore the posterior distribution. Specifically, the funnel geometry arises due to the strong coupling between the scale parameter *τ* and the weights *θ*, where small values of *τ* compress the posterior distribution into a narrow region, and large values of *τ* allow for wider exploration. This creates a highly anisotropic posterior, with steep gradients near the origin and flat gradients in the outer regions. Thereby, MCMC samplers, particularly gradient-based algorithms like the No-U-Turn Sampler (NUTS), struggle to traverse these regions efficiently. The steep gradients require very small step sizes to navigate, slowing the sampling process, while the flat gradients in the outer regions lead to poor exploration of the parameter space. This inefficiency not only prolongs computation but also results in poor mixing and limited exploration of the posterior, thereby affecting the accuracy and reliability of the parameter estimates. The funnel structure ultimately makes the posterior difficult to sample from, especially when the data strongly inform the weights *θ* but provide weak information about the variance parameter *τ*, further exacerbating the challenges in convergence and computational burden. Consequently, the sampling process suffers from high variability and poor mixing, which not only slows computational performance but also contributes to the convergence issues observed for *τ*. Specifically, the inability to adequately explore the posterior space exacerbates the instability in estimating the spread of significant weights, further highlighting the challenges in distinguishing between small but relevant features and random noise.

To address this issue, non-centered parameterization offers a promising solution by redefining *θ* in terms of a standard normal variable 𝒵 ∼ 𝒩 (0, 1) such that *θ* = *τ* · 𝒵. This transformation decouples the dependence between *θ* and *τ*, effectively removing the funnel-shaped posterior geometry and simplifying the sampling process. By enabling more uniform exploration of the parameter space, non-centered parameterization improves the efficiency and stability of MCMC algorithms, reducing the runtime and enhancing the convergence of *τ*. Applying this adjustment in future implementations of the Spike-and-Slab method could not only mitigate computational inefficiencies but also improve the reliability of parameter estimates, making the method more robust and scalable for larger datasets [31].

#### Bayesian LASSO

The Bayesian LASSO implementation utilized Hamiltonian Monte Carlo sampling with the No-U-Turn Sampler, employing 2 parallel chains with 2,000 post-warm-up draws each. The model demonstrated robust convergence across all parameters, with Gelman-Rubin statistics 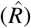 ranging from 1.000 to 1.003, substantially below the conventional threshold of 1.1. The global shrinkage parameter *τ* exhibited particularly strong convergence 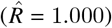, with posterior mean 0.711 (97% HDI: [0.603, 0.820]) and high sampling efficiency (ESS bulk = 2933.0, ESS tail = 2893.0). The model intercept showed similar stability (ESS bulk = 6257.0, posterior mean -2.720, 97% HDI: [-2.958, -2.478]). The trace plots (Figure 8) demonstrate excellent mixing behavior and rapid convergence. The left panels reveal symmetric, well-formed posterior densities for both *τ* and the intercept, indicating proper posterior exploration. The right panels show stable trajectories without persistent trends or anomalous patterns, with the two chains exhibiting appropriate overlap and stationary behavior throughout the sampling period. The smooth transitions between states and consistent exploration of the posterior space support the validity of our sampling approach. From 120 initial predictors, the model identified 90 features exceeding the 75th percentile importance threshold. The posterior estimates revealed a clear separation in feature importance, with the three most influential predictors demonstrating substantial standardized effects: Feature 5 (-4.876, importance score 28.684), Feature 3 (3.777, importance score 24.924), and Feature 4 (-2.177, importance score 21.136). The marked decline in coefficient magnitudes beyond these top predictors (next largest —coefficient— = 0.413) suggests natural sparsity in the feature space, validating the selective shrinkage properties of the Bayesian LASSO framework. The computational demands were considerable (runtime: 11 hours 16 minutes) due to the large sample size (N > 23,000), but the excellent convergence diagnostics and low Monte Carlo standard errors (all MCSE < 0.002) justify this investment for ensuring reliable posterior inference.

**Figure 8.**
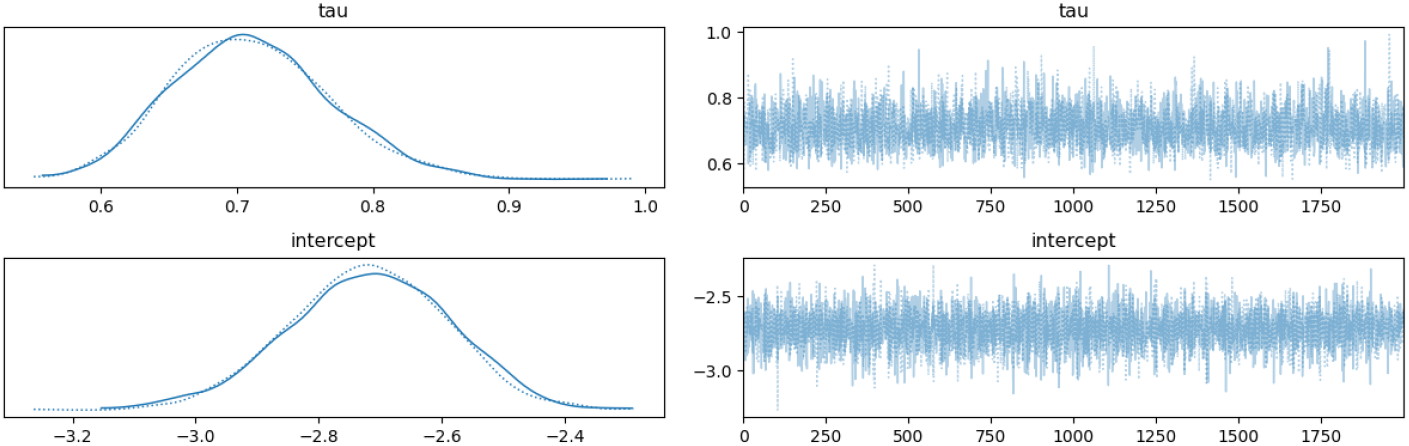
MCMC diagnostic plots for the Bayesian LASSO model. Trace plots for model parameters (*τ* and intercept) showing mixing behavior across two chains. Left panels show smoothed posterior densities; right panels show detailed sampling trajectories over iterations.

#### Standard LASSO

In the real-world dataset analysis, the LASSO method successfully shrank 14,454 out of 14,976 weights to zero, resulting in the removal of 34 out of 117 input features. This corresponds to retaining approximately 70.94% of the original features, demonstrating LASSO’s ability to enforce sparsity not only at the weight level but also at the feature level, in contrast to the simulated results where no features were removed. The feature importance scores, shown in Figure 9, reveal that a small subset of features dominated the predictive performance, with the remaining features exhibiting low importance. This result highlights LASSO’s effectiveness in discarding negligible predictors while retaining the most informative ones.

**Figure 9.**
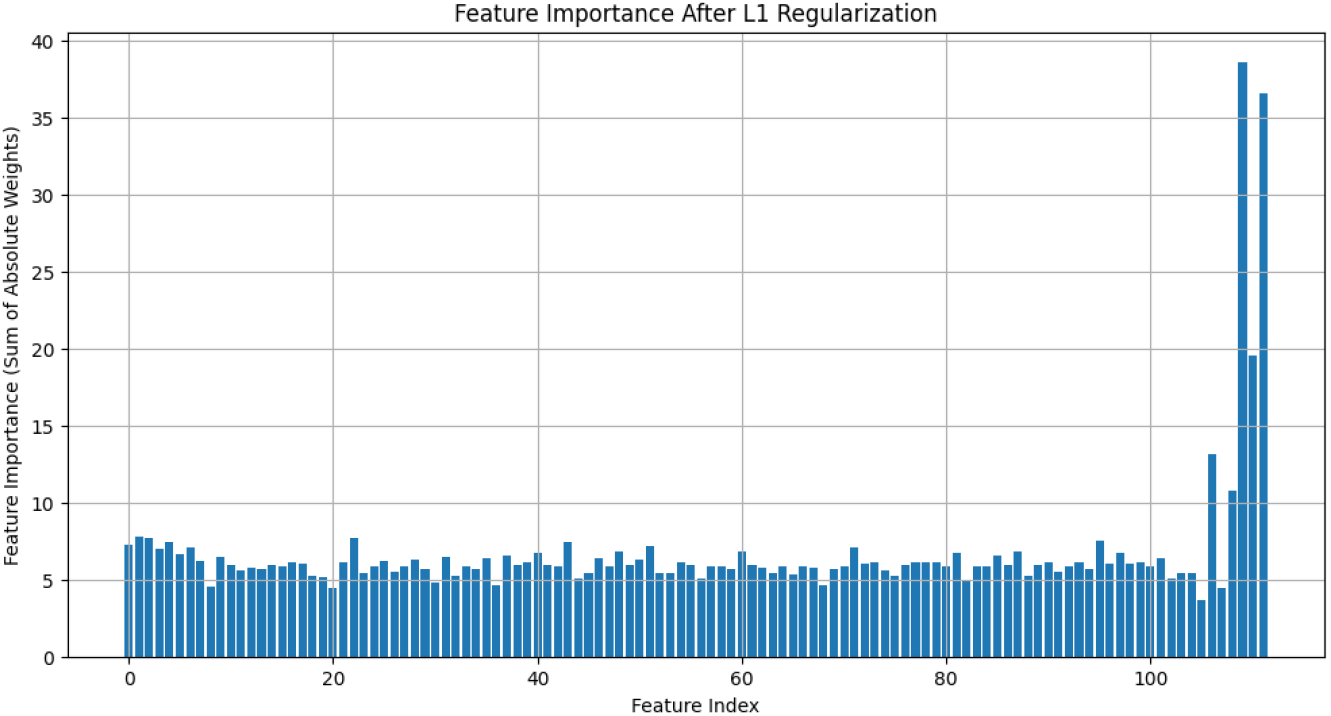
Feature importance after LASSO regularization in the real-world dataset.

The standard LASSO model achieved a test accuracy of 0.8074, which, while competitive, was slightly inferior to the Bayesian LASSO (0.823) and spike-and-slab (0.822) methods. The observed performance gap underscores LASSO’s limitations in capturing model uncertainty, which is a notable advantage of Bayesian frameworks.

These findings reinforce the utility of *L*_1_-regularization for enforcing sparsity and simplifying models, particularly in real-world datasets with more variability and noise compared to simulated data.

### Model output

Table 1 provides a comprehensive comparative analysis of performance metrics for the four methods—Spike-and-Slab, Bayesian LASSO, LASSO, and All Features (inclusion of all features without selection techniques)—highlighting their relative effectiveness in predictive modeling. The All Features method achieves the highest precision (78.20%), indicating strong performance in minimizing false positives. However, its recall (39.33%) is relatively moderate compared to Spike (44.96%), which demonstrates the highest recall among the methods. The Spike model also achieves the highest F1-score (55.46%), reflecting its ability to balance precision and recall effectively. Bayesian LASSO shows competitive performance, achieving strong precision (75.93%) and a moderate recall (40.67%), while LASSO demonstrates the weakest performance in recall (32.52%) and F1-score (45.33%).

**Table 1:** Performance Metrics and Pairwise Percentage Improvements Across Methods.

| Performance Metrics |  |  |  |  |
| --- | --- | --- | --- | --- |
| Method | Precision (%) | Recall (%) | F1 (%) | AUC (%) |
| SPIKE | 72.35 | 44.96 | 55.46 | 85.31 |
| Bayesian LASSO | 75.93 | 40.67 | 52.97 | 85.57 |
| LASSO | 74.79 | 32.52 | 45.33 | 83.16 |
| All Features | 78.20 | 39.33 | 52.34 | 86.17 |
| Pairwise Percentage Improvements |  |  |  |  |
| Comparison | Precision (%) | Recall (%) | F1 (%) | AUC (%) |
| SPIKE vs LASSO | -3.26 | 38.33 | 22.38 | 2.58 |
| Bayesian LASSO vs LASSO | 1.52 | 24.98 | 16.27 | 2.90 |
| SPIKE vs Bayesian LASSO | -4.72 | 10.58 | 4.70 | -0.30 |
| SPIKE vs All Features | -7.46 | 14.30 | 6.00 | -1.00 |

The AUC metric, which measures the overall discriminative ability of the models, is highest for All Features (86.17%), closely followed by Bayesian LASSO (85.57%) and Spike (85.31%). While the All Features method excels in AUC and precision, the inclusion of all features introduces noise that diminishes the interpretability and robustness of the model, as indicated by its lower F1-score. In contrast, the Spike model’s competitive AUC and superior F1-score make it a robust alternative that balances predictive performance and feature selection.

The pairwise comparisons highlight the performance trade-offs between the methods. When comparing Spike-and-Slab to LASSO, the Spike-and-Slab model exhibits a substantial improvement in recall (38.33%) and F1-score (22.38%), underscoring its ability to identify true positives effectively. However, this improvement comes with a minor trade-off in precision (-3.27%). Similarly, Bayesian LASSO shows notable improvements over LASSO, with a 24.98% increase in recall and a 16.27% improvement in F1-score, while achieving a modest gain in precision (1.52%).

A comparison between Spike-and-Slab and Bayesian LASSO reveals that Spike-and-Slab outperforms Bayesian LASSO in recall (10.58%) and F1-score (4.70%), emphasizing its effectiveness in identifying true positives. Despite a slight reduction in precision (-4.72%), the Spike-and-Slab model’s superior recall and F1-score demonstrate its ability to minimize false negatives without significantly compromising precision. Compared to the All Features method, Spike achieves a 14.30% improvement in recall and a 6.00% increase in F1-score, further underscoring its robust performance. However, Spike-and-Slab trails behind All Features in precision (-7.46%) and AUC (-1.00%), indicating that the All Features method may excel in scenarios prioritizing false positive minimization.

Among the methods, the Spike-and-Slab model emerges as the best overall performer, achieving the highest F1-score and recall, which are critical metrics for balanced classification tasks. While including all features achieves the highest precision and AUC, its lower recall highlights its susceptibility to false negatives, limiting its utility in scenarios where recall is vital. Bayesian LASSO provides a competitive alternative with balanced improvements over LASSO but does not surpass Spike in overall performance. The Spike model’s ability to combine effective feature selection with superior recall and F1-score underscores its robustness in optimizing predictions. These results highlight Spike and Slab feature selection as the most effective and balanced method for real-world data analysis, ensuring accurate and interpretable outcomes.

### Clinical Significance

Our comparative effectiveness analysis employed multiple feature selection approaches within a causal inference framework to evaluate treatment effects, with all methods revealing significant differences between vasopressors (Figure 10). The Spike-and-Slab and LASSO methods demonstrated comparable precision in effect estimates, with LASSO offering computational advantages while maintaining similar confidence interval widths, particularly in the vasopressin versus phenylephrine comparison (Figure 10A). Across all methodological approaches, vasopressin consistently demonstrated superior effectiveness compared to both alternative agents, supporting strong causal inference regarding treatment effects. Our optimal, Spike-and-Slab estimates indicated that vasopressin treatment was associated with significantly better outcomes compared to norepinephrine (ATE = 0.134, 95% CI [0.120, 0.152], p < 0.001) and phenylephrine (ATE = 0.173, 95% CI [0.156, 0.191], p < 0.001). The magnitude of these causal effects remained remarkably stable across different feature selection methods, with mean estimates varying by less than 0.01 between methods, strengthening causal interpretations of our findings. Phenylephrine consistently showed inferior outcomes compared to norepinephrine across all analyses, with the LASSO estimating this causal effect at ATE = -0.040 (95% CI [-0.048, -0.031], p < 0.001). This finding was particularly notable for its consistency across methods, with all approaches yielding similar point estimates and overlapping confidence intervals, supporting robust causal inference (Figure 10A). The comparative analysis revealed a clear hierarchical pattern in treatment effectiveness (Figure 10B). Vasopressin emerged as the most effective agent, with estimates suggesting an absolute risk reduction of approximately 13-17 percentage points compared to alternative agents. These findings represent clinically meaningful treatment effects that remain consistent across multiple analytical approaches, strengthening their validity for clinical decision-making.

**Figure 10.**
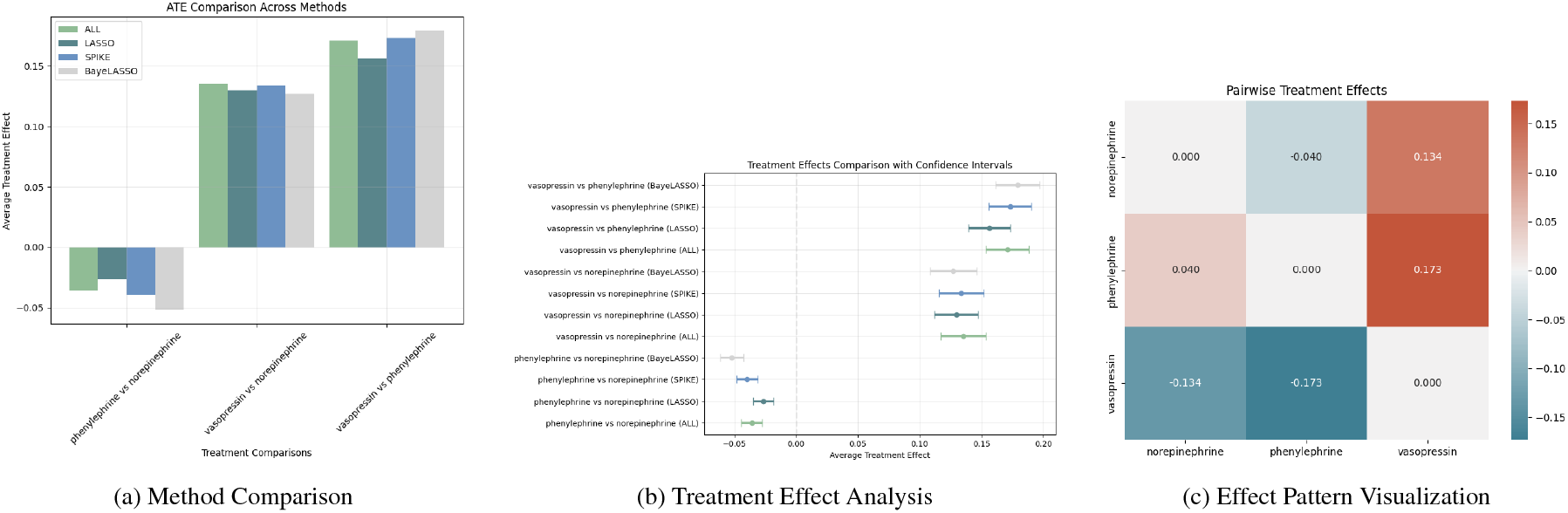
Comparative analysis of vasopressor treatment effects using different feature selection methods. This figure presents a comprehensive evaluation of treatment effectiveness across multiple analytical approaches. (a) Bar plot comparing average treatment effects (ATEs) across four feature selection methods (ALL, LASSO, spike-and-slab, and Bayesian LASSO), demonstrating consistency in effect direction and magnitude across methodological approaches. (b) Forest plot displaying pairwise ATEs with 95% confidence intervals for each feature selection method, where positive values indicate superior effectiveness of the second listed treatment. The vertical dashed line at zero represents no treatment difference, with points to the right indicating better outcomes for the comparator treatment. (c) Heatmap visualization derived from the spike-and-slab model (best performing method) showing the pattern of treatment effects, where blue indicates better outcomes (lower mortality) and red indicates worse outcomes (higher mortality). The color intensity corresponds to the magnitude of the treatment effect difference

## Discussion

Recent studies indicate substantial variation in mortality rates (15-60%) among shock patients receiving vasopressors [32, 33], underscoring the critical need for more sophisticated treatment selection methods. This variation reflects three key challenges in vasopressor research: identifying clinically meaningful predictors from hundreds of variables [34], capturing nonlinear treatment effect heterogeneity across patient subgroups [35], and addressing confounding by indication where both treatment assignment and outcomes are influenced by unmeasured severity indicators [36].

Our large-scale observational study provides important causal insights into these challenges through several methodological innovations. By implementing Bayesian spike and slab priors within a causal inference framework, we achieved optimal bias control while enabling more robust feature selection to identify key effect modifiers. The Bayesian neural network framework captures complex nonlinear relationships while quantifying prediction uncertainty. Notably, while the Spike-and-Slab method demonstrated superior bias control, the LASSO approach provided comparable accuracy with enhanced computational efficiency, offering a practical alternative for large-scale analyses.

These methodological advances yielded important clinical insights. The superior performance of vasopressin aligns with foundational experimental work by Minneci (2004), who demonstrated beneficial effects of both vasopressin and norepinephrine in septic shock models [37]. Our results extend these findings to establish causal relationships in a real-world clinical population, showing that vasopressin’s advantages persist across a heterogeneous patient population (ATE = 0.134 vs norepinephrine, p < 0.001).

The significant inferiority of phenylephrine compared to norepinephrine (ATE = -0.040, p < 0.001) corresponds with findings from Wang (2018) [38]. Our causal inference framework strengthens these previous associations by establishing clearer evidence of direct treatment effects. The consistency of these effects across multiple analytical methods particularly reinforces the robustness of our causal conclusions. Our causal inference analysis provides robust statistical evidence supporting the existing clinical framework for vasopressor use in shock, demonstrating that modern Bayesian and machine learning methods (Spike-and-Slab and LASSO) can effectively quantify treatment effects that align with established clinical practice recommendations [39].

Our methodological approach offers several strengths in establishing causal relationships. The instrumental variable approach using physician prescribing preferences helps address confounding by indication, while our comparison of feature selection methods demonstrates the robustness of our causal estimates. The Spike-and-Slab method’s superior bias control, coupled with LASSO’s computational efficiency and comparable accuracy, provides a practical framework for future causal inference studies in critical care. However, our study still limited by several important features. First, while physician prescribing preferences serve as our instrumental variable, we acknowledge potential challenges to this approach. Physicians with certain prescribing patterns may systematically treat patients with different severity levels, potentially compromising the exclusion restriction. Additionally, variations in prescribing patterns might correlate with other aspects of care delivery, though we attempted to minimize this by adjusting for physician characteristics and hospital-level factors. Second, our study relies on data from a single center (MIMIC-IV database from Beth Israel Deaconess Medical Center), which may limit generalizability. While our methodological framework is broadly applicable across different settings, the specific treatment effects we observed may vary in institutions with different patient populations, clinical protocols, or prescribing practices. Institutional factors such as ICU staffing models, hospital protocols, and available resources could influence both treatment selection and outcomes. Third, despite our sophisticated methodological approach, unmeasured confounding may persist. This is particularly relevant in critical care settings, where subtle clinical factors might influence both treatment selection and outcomes.

Future research should focus on external validation of these causal relationships in multi-center studies and investigating potential effect modifiers to identify patient subgroups who might particularly benefit from specific vasopressors. The methodological framework developed here, particularly the efficient LASSO approach, could be extended to other clinical scenarios requiring robust causal inference with high-dimensional covariates and unmeasured confounding. Prospective trials comparing different timing strategies for vasopressin initiation may be particularly valuable given our findings regarding its effectiveness. The generalizability of our analytical framework to other settings represents a key strength - while our specific findings reflect one institution’s experience, the methods we’ve developed can be readily applied to similar analyses in different healthcare settings, potentially yielding insights specific to local populations and practice patterns.

Our findings have immediate implications for clinical practice while acknowledging the need for validation in diverse settings. The robust causal evidence supporting vasopressin’s superiority suggests that current prescribing patterns may warrant reconsideration, though individual patient factors and local institutional practices should inform treatment decisions. This methodological framework and implementation code are publicly available (https://github.com/Bma0828/BayesianMultiValuedEHR.git), facilitating reproduction and extension of these analyses.

## Data Availability

All data produced in the present study are available upon reasonable request to the authors

https://github.com/Bma0828/BayesianMultiValuedEHR.git

## Acknowledgements

We would like to acknowledge the distinct contributions of our team members. Yunzhe Qian led the overall research design and methodology development, with primary contributions including the conceptualization of the entire Bayesian instrumental variable framework, implementation of the Bayesian LASSO feature selection method, and assisted with the neural network outcome regression analysis. She was also responsible for data acquisition and preprocessing from the MIMIC-IV database, conducting the primary statistical analyses, and leading the manuscript preparation. Bowen Ma contributed to the implementation of the multinomial logistic treatment regression model, spike-and-slab feature selection analysis, standard LASSO feature selection analysis, and development of the neural network instrumental variable approach. Both authors participated in the refinement of the methodology, manuscript, and slides. We also thank the PhysioNet team for providing access to the MIMIC-IV database, which made this research possible.

## References

[1] Alistair EW Johnson, Lucas Bulgarelli, Lu Shen, Alvin Gayles, Ayad Shammout, Steven Horng, Tom J Pollard, Sicheng Hao, Benjamin Moody, Brian Gow, et al. Mimic-iv, a freely accessible electronic health record dataset. Scientific data, 10(1):1, 2023.

[2] M Alan Brookhart, Philip S Wang, Daniel H Solomon, and Sebastian Schneeweiss. Evaluating short-term drug effects using a physician-specific prescribing preference as an instrumental variable. Epidemiology, 17(3):268–275, 2006.

[3] Jeremy A Rassen, M Alan Brookhart, Robert J Glynn, Murray A Mittleman, and Sebastian Schneeweiss. Instrumental variables ii: instrumental variable application—in 25 variations, the physician prescribing preference generally was strong and reduced covariate imbalance. Journal of clinical epidemiology, 62(12):1233–1241, 2009.

[4] M Alan Brookhart, Jeremy A Rassen, Philip S Wang, Colin Dormuth, Helen Mogun, and Sebastian Schneeweiss. Evaluating the validity of an instrumental variable study of neuroleptics: can between-physician differences in prescribing patterns be used to estimate treatment effects? Medical care, 45(10):S116–S122, 2007.

[5] Jessica Gronsbell, Jessica Minnier, Sheng Yu, Katherine Liao, and Tianxi Cai. Automated feature selection of predictors in electronic medical records data. Biometrics, 75(1):268–277, 2019.

[6] Zheming Zuo, Jie Li, Han Xu, and Noura Al Moubayed. Curvature-based feature selection with application in classifying electronic health records. Technological Forecasting and Social Change, 173:121127, 2021.

[7] Rishi J Desai, Mufaddal Mahesri, Younathan Abdia, Julie Barberio, Angela Tong, Dongmu Zhang, Panagiotis Mavros, Seoyoung C Kim, and Jessica M Franklin. Association of osteoporosis medication use after hip fracture with prevention of subsequent nonvertebral fractures: an instrumental variable analysis. JAMA network open, 1(3):e180826–e180826, 2018.

[8] Nicolas Bastardoz, Michael J Matthews, Gwendolin B Sajons, Tyler Ransom, Thomas K Kelemen, and Samuel H Matthews. Instrumental variables estimation: Assumptions, pitfalls, and guidelines. The Leadership Quarterly, 34(1):101673, 2023.

[9] Jesse A Columbo, Pablo Martinez-Camblor, Todd A MacKenzie, Douglas O Staiger, Ravinder Kang, Philip P Goodney, and A James O’Malley. Comparing long-term mortality after carotid endarterectomy vs carotid stenting using a novel instrumental variable method for risk adjustment in observational time-to-event data. JAMA network open, 1(5):e181676–e181676, 2018.

[10] Brandon Lu, Sasha Thomson, Scott Blommaert, Mina Tadrous, Craig C Earle, and Kelvin KW Chan. Use of instrumental variable analyses for evaluating comparative effectiveness in empirical applications of oncology: A systematic review. Journal of Clinical Oncology, 41(13):2362–2371, 2023.

[11] M Alan Brookhart, Jeremy A Rassen, and Sebastian Schneeweiss. Instrumental variable methods in comparative safety and effectiveness research. Pharmacoepidemiology and drug safety, 19(6):537–554, 2010.

[12] Guido W Imbens. The role of the propensity score in estimating dose-response functions. Biometrika, 87(3):706–710, 2000.

[13] Kosuke Imai and David A Van Dyk. Causal inference with general treatment regimes: Generalizing the propensity score. Journal of the American Statistical Association, 99(467):854–866, 2004.

[14] Ariel Linden, S Derya Uysal, Andrew Ryan, and John L Adams. Estimating causal effects for multivalued treatments: a comparison of approaches. Statistics in Medicine, 35(4):534–552, 2016.

[15] Daniel F McCaffrey, Beth Ann Griffin, Daniel Almirall, Mary Ellen Slaughter, Rajeev Ramchand, and Lane F Burgette. A tutorial on propensity score estimation for multiple treatments using generalized boosted models. Statistics in medicine, 32(19):3388–3414, 2013.

[16] Jason Hartford, Greg Lewis, Kevin Leyton-Brown, and Matt Taddy. Counterfactual prediction with deep instrumental variables networks. arXiv preprint arXiv:1612.09596, 2016.

[17] Zihang Lu and Wendy Lou. Bayesian approaches to variable selection: a comparative study from practical perspectives. The International Journal of Biostatistics, 18(1):83–108, 2022.

[18] Neil M Davies, David Gunnell, Kyla H Thomas, Chris Metcalfe, Frank Windmeijer, and Richard M Martin. Physicians’ prescribing preferences were a potential instrument for patients’ actual prescriptions of antidepressants. Journal of clinical epidemiology, 66(12):1386–1396, 2013.

[19] Ernst R Berndt, Robert S Gibbons, Anton Kolotilin, and Anna Levine Taub. The heterogeneity of concentrated prescribing behavior: Theory and evidence from antipsychotics. Journal of health economics, 40:26–39, 2015.

[20] Yan Tang, Chung-Chou H Chang, Judith R Lave, Walid F Gellad, Haiden A Huskamp, and Julie M Donohue. Patient, physician and organizational influences on variation in antipsychotic prescribing behavior. The journal of mental health policy and economics, 19(1):45, 2016.

[21] Michael Keane and Timothy Neal. Instrument strength in iv estimation and inference: A guide to theory and practice. Journal of Econometrics, 235(2):1625–1653, 2023.

[22] John Salvatier, Thomas V Wiecki, and Christopher Fonnesbeck. Probabilistic programming in python using pymc3, 2016.

[23] Trevor Park and George Casella. The bayesian lasso. Journal of the american statistical association, 103(482):681–686, 2008.

[24] Andrew Gelman. Prior distributions for variance parameters in hierarchical models (comment on article by browne and draper). 2006.

[25] Nicholas G Polson and James G Scott. On the half-cauchy prior for a global scale parameter. Bayesian Analysis, 7(4):887–902, 2012.

[26] TensorFlow Developers. Tensorflow keras regularizers documentation. TensorFlow Documentation, 2024. https://www.tensorflow.org/api_docs/python/tf/keras/regularizers/L1.

[27] Aki Vehtari, Andrew Gelman, Daniel Simpson, Bob Carpenter, and Paul-Christian Bürkner. Rank-normalization, folding, and localization: An improved 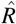 for assessing convergence of MCMC. Bayesian Analysis, 16(2):667–718, 2021.

[28] Andrew Gelman and Donald B Rubin. Inference from iterative simulation using multiple sequences. Statistical Science, 7(4):457–472, 1992.

[29] Jonah Gabry, Daniel Simpson, Aki Vehtari, Michael Betancourt, and Andrew Gelman. Visualization in Bayesian workflow. Journal of the Royal Statistical Society: Series A, 182(2):389–402, 2019.

[30] Radford M. Neal. Slice sampling. The Annals of Statistics, 31(3):705–767, 2003.

[31] Stan Development Team. Efficiency tuning for Stan programs: Reparameterization and other techniques, 2024. Accessed December 2024.

[32] Alexandria C Rydz, Jessica L Elefritz, Megan Conroy, Kathryn A Disney, Christopher J Miller, Kyle Porter, and Bruce A Doepker. Early initiation of vasopressin reduces organ failure and mortality in septic shock. Shock, 58(4):269–274, 2022.

[33] Russel J Roberts, Todd A Miano, Drayton A Hammond, Gourang P Patel, Jen-Ting Chen, Kristy M Phillips, Natasha Lopez, Kianoush Kashani, Nida Qadir, Charles B Cairns, et al. Evaluation of vasopressor exposure and mortality in patients with septic shock. Critical care medicine, 48(10):1445–1453, 2020.

[34] Gertraud Malsiner-Walli and Helga Wagner. Comparing spike and slab priors for bayesian variable selection. arXiv preprint arXiv:1812.07259, 2018.

[35] Yarin Gal and Zoubin Ghahramani. Bayesian convolutional neural networks with bernoulli approximate variational inference. arXiv preprint arXiv:1506.02158v6, 2016.

[36] Demetrios N Kyriacou and Roger J Lewis. Confounding by indication in clinical research. Jama, 316(17):1818–1819, 2016.

[37] PC Minneci, KJ Deans, SM Banks, R Costello, and C Natanson. Differing effects of epinephrine, norepinephrine, and vasopressin on survival in a canine model of septic shock. American Journal of Physiology-Heart and Circulatory Physiology, 287(6):H2545–H2554, 2004.

[38] Xian Wang, Min Mao, Shijiang Liu, Shiqin Xu, and Jie Yang. The efficacy and safety of norepinephrine and its feasibility as a replacement for phenylephrine to adjust maternal blood pressure during caesarean section under spinal anesthesia. BioMed Research International, 2018, 2018.

[39] Matthew S Heavner, Gautam V Ramani, and Kristina R Claeys. Angiotensin ii and vasopressin for vasodilatory shock: A critical appraisal of catecholamine-sparing strategies. Critical Care Explorations, 2(9):e0181, 2020.

